# Labor market participation and depression during the COVID-19 epidemic among young adults (18 to 30 years): a nationally representative study in France

**DOI:** 10.1101/2022.03.25.22272948

**Authors:** Maria Melchior, Aline-Marie Florence, Camille Davisse-Paturet, Bruno Falissard, Cédric Galéra, Jean-Baptiste Hazo, Cécile Vuillermoz, Josiane Warszawski, Fallou Dione, Alexandra Rouquette, the EPICOV study group

## Abstract

**Objective:** To examine the relationship between young adults’ labor force participation and depression in the context of the COVID-19 pandemic.

**Design, Setting, Participants:** Data come from the nationally-representative EPICOV cohort study set up in France, and were collected in 2020 and 2021 (3 waves of online or telephone interviews) among 2217 participants aged 18-30 years. Participants with prior mental health disorder (n=50) were excluded from the statistical analyses.

**Results:** Using Generalized Estimating Equation (GEE) models controlled for participants’ socio-demographic and health characteristics and weighted to be nationally-representative, we found that compared to young adults who were employed, those who were studying or unemployed were significantly more likely to experience depression assessed using the PHQ-9 (multivariate ORs respectively: OR: 1.29, 95% CI 1.05-1.60 and OR: 1.50, 1.13-1.99). Stratifying the analyses by age, we observed than unemployment was more strongly associated with depression among participants 25-30 years than among those who were 18-24 years (multivariate ORs respectively 1.78, 95% CI 1.17-2.71 and 1.41, 95% CI 0.96-2.09). Being out of the labor force was, to the contrary, more significantly associated with depression among participants 18-24 years (multivariate OR: 1.71, 95% CI 1.04-2.82, vs. 1.00, 95% CI 0.53-1.87 among participants 25-30 years). Stratifying the analyses by sex, we found no significant differences in the relationships between labor market characteristics and depression (compared to participants who were employed, multivariate ORs associated with being a student: men: 1.33, 95% CI 1.01-1.76; women: 1.19, 95% CI 0.85-1.67, multivariate ORs associated with being unemployed: men: 1.60, 95% CI 1.04-2.45; women: 1.47, 95% CI 1.01-2.15).

**Conclusions and relevance:** Our study shows that in addition to students, young adults who are unemployed also experience elevated levels of depression in the context of the COVID-19 pandemic. These two groups should be the focus of specific attention in terms of prevention and mental health treatment. Supporting employment could also be a propitious way of reducing the burden of the Covid-19 pandemic on the mental health of young adults.

**Key Points:** *Question:* Is labor force participation associated with young adults’ likelihood of depression during the COVID-19 pandemic?

*Findings:* In a nationally-representative cohort study in France, compared to young adults who are employed, those who are studying or experience unemployment had elevated odds of depression in 2020 and 2021.

*Meaning:* Young people are experiencing the highest burden of mental health problems in the context of the COVID-19 epidemic – our study implies that those who are studying or are unemployed are at especially high risk and should be the focus of attention in terms of prevention and treatment.

## Introduction

The COVID-19 pandemic has had a major impact on worldwide population mental health, still unresolved today (1). In particular, there is evidence of significant increases in the prevalence of major depression (up to approximately 27%) and anxiety disorders (up to approximately 25%), respectively leading to an estimated 50 and 76 million new cases globally (1). In the general population, factors associated with a deterioration in mental health include female sex, age under 35 years, loneliness and low income (2). In addition, several studies have found that university students experienced especially large increases in the prevalence of depression and anxiety disorders, with approximately a third suffering from either condition (3-5). However, most of these studies were conducted in China (3-5) where the spread of COVID-19 was limited after 2019 and accompanied by strict preventive measures, or in the United States (3) where to the contrary, the toll of the COVID-19 epidemic has been high and distance teaching was implemented for extended periods of time. Data from additional countries can help better understand the extent to which students are a high-risk group in terms of mental health during in the context of the current sanitary crisis.

Moreover, young people are disproportionately likely to experience job instability and unemployment, which increased during the course of the COVID-19 epidemic and have been identified as risk factors of depression and anxiety (6-10). The association between unemployment and mental health has previously been observed and is considered to be partly bidirectional (11), indicating that it is essential to consider preexisting mental health difficulties in assessing changes in psychological distress during the course of the COVID-19 epidemic.

France is characterized by high levels of youth unemployment compared to other OECD countries: 18.5% among 15-24 year-olds in 2021, as compared with 11.5% on average (12). Additionally, a high proportion of youths are Not in Employment, Education or Training (NEET): 19% of 20-24 year-olds as compared to 15.5% on average (13). Youth unemployment appears to have increased to about 21% in 2020 before decreasing to about 18% by the end of 2021 (14), suggesting heightened pressure on young adults attempting to find employment and making France a relevant context to study relationships with mental health.

Moreover, France is among countries which were hit especially hard by the COVID-19 pandemic, particularly in 2020 and 2021. By March 2022, the COVID-19 epidemic has caused the death of over 140 000 persons (https://coronavirus.jhu.edu/region/france) and resulted in a strict national lockdown from March to May 2020, followed by two mitigated lockdowns from November 2020 to January 2021 and from April to June 2021. Between these periods, a curfew was implemented, and social and educational activities were restricted. According to a nationally representative survey of the French population conducted by the French Public Health Agency, the prevalence of symptoms of depression among 18-24 year-olds was estimated to be 16% in the Spring 2020 and 22% in January 2022, while respectively 33% and 43% suffered from symptoms of anxiety, that is double the rates observed prior to the COVID-19 epidemic (15). Similarly, a study conducted by the French Ministry of Health estimated, that in May and November 2020, respectively 22 and 19 % of 15-24 years old experienced symptoms of depression while the prevalence prior to the COVID-19 pandemic was estimated to be 10% (16).

The aim of our study was to test the association between labor market participation and depression among young adults during the course of the COVID-19 epidemic. In particular, we compared young adults who were employed to those who were studying or unemployed, while accounting for sociodemographic and health factors potentially associated with depressive symptoms, including history of mental health problems prior to the COVID-19 epidemic.

## Methods

### Study design

Data come from the EPICOV study, a French nationally-representative cohort designed to assess the main characteristics of the COVID-19 pandemic on the population of France as well as its impact in terms of sociodemographic and health outcomes (17). As previously described, participants (≥ 15 years of age, residing in mainland France or three out of five overseas territories) were randomly selected from the national tax database (FIDELI). FIDELI covers 96.4% of the population living in France, providing postal addresses for all individuals, and an e-mail address or telephone number for 83%. Sampling was stratified on two characteristics: residential area (‘département’ - equivalent to a county), and area-level poverty defined as 60% of the median household income per capita. This sampling frame was designed to ensure overrepresentation of less densely populated and more socioeconomically disadvantaged areas. Individuals living in residential care or prison were not invited to participate in the study.

All potential participants were contacted by post, e-mail and text messages (SMS), with up to seven reminders. Self-computer-assisted-web interviews (CAWI) were conducted with 80% of study participants. The remaining 20% were randomly assigned to CAWI or computer-assisted-telephone interviews (CATI).

The first wave of data collection took place in the Spring 2020 (02/05/2020 – 12/06/2020) at the end of the first national COVID-19-related lockdown; the second in the Fall 2020 (26/10/2020 – 14/12/2020), during the second national COVID-19-related lockdown; the third in the Summer 2021 (24/06/2021 – 09/08/2021) after the end of the third national COVID-19-related lockdown.

### Study population

Initially, 371,000 persons were randomly selected and 134,391 completed the first EPICOV study questionnaire (36.4% participation). Measures of mental health were exclusively included in an extended version of the study questionnaire completed by 10% of the study sample followed-up longitudinally (n= 14,237 persons in the first study wave, n=12,519 in the second and n=10,780 in the third study wave). Since our aim was to study determinants of depression among young adults, statistical analyses were restricted to participants aged 18-30 years who had at least one measure of depression during the course of follow-up (n= 2,271 persons in the first, n=1,966 in the second and n=1,615 in the third study wave). Participants’ aged 15-17 years at the time of the study were excluded because 95,57% were students, therefore there was no variability in labor force participation in this group. Additionally, to limit the influence of preexisting mental health difficulties on our results, we excluded study participants who had a prior history of depression or anxiety which limited their daily activities (n=50) (**Figure 1**). In our study sample, 74.2% of participants completed the study questionnaire by CAWI and 25.8% by CATI.

**Figure 1.**
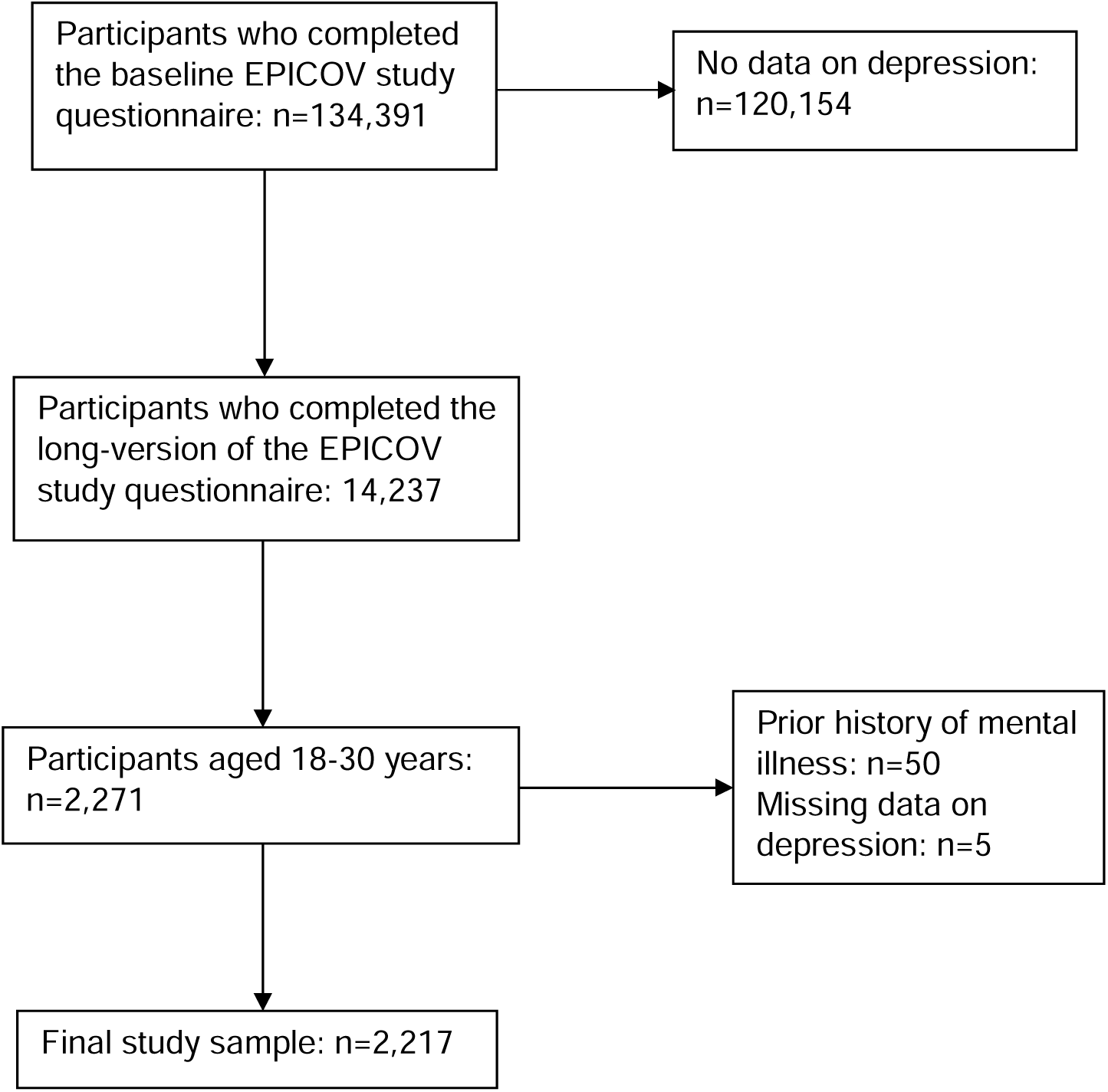
Flowchart showing the selection of EPICOV study participants aged 18-30 years and with available depression data.

### Study outcome

The primary outcome in our study was depression over the two weeks preceding the assessment, measured by the Patient Health Questionnaire – 9 items (PHQ-9) score (18). Following previously published guidelines, participants with a score >= 10 were considered to have moderate to severe depression and were considered as cases.

### Labor market participation

Participants’ labor market participation was assessed at each study wave and treated as a time-varying exposure. We distinguished participants who reported that they were employed, studying, unemployed or out of the labor force (that is neither employed, unemployed, nor studying). Participants who reported studying and working were considered to be primarily students.

### Covariates

Based on prior evidence, we selected several socio-demographic and health characteristics as possible confounding variables of the association between labor market participation and depression. Several characteristics were measured at study baseline: sex (female vs. male), age (25-30 vs. 18-24 years), urbanicity (rural area, <100 000 residents, ≥100 000 residents, Paris and suburbs), lone living (yes vs. no), having children (yes vs. no), and presence of a chronic disease (yes vs. no, the most frequent diseases reported were asthma - 4.6%, physical disability - 1.1%, gastro-intestinal disorder - 0.9%, musculo-skeletal disorder - 0.9%, respiratory disorder - 0.6%, diabetes - 0.5%). Other characteristics were studied as time-dependent variables: perceived financial situation (ascertained by the following question: ‘Do you consider your financial situation to be - comfortable, suitable, tight, difficult?’), romantic relationship (yes vs. no), outings in preceding week (<once, 2-5 times, >5 times), and experience of COVID-19 symptoms (yes vs. no, one or more symptoms among cough, fever, dyspnea, or ageusia, dysgeusia or anosmia).

### Statistical analyses

Reweighting and marginal calibrations were used to correct for non-participating bias and to take into account the effect of the means of data collection (Internet or telephone). To test the association between young adults’ labor market participation and depression during the COVID-19 epidemic we proceeded as follows. First, associations were tested in bivariate Generalized Estimating Equations (GEE)(19) regression models weighted for study weights. Second, a multivariate GEE regression model was implemented including the study wave and all covariates associated with the study outcome with a p-value <0.20 in bivariate models. In subsequent analyses, we repeated the analyses stratified on age (18-24 / 25-30 years), 25 being the age when youths in France cannot anymore be claimed as dependents by their parents. Additionally, we tested statistical interactions between participants’ labor market participation and sex. Missing data on covariates (maximum 37%) were imputed using multiple imputations under a missing at random hypothesis (20). Data management and descriptive analyses were performed using R 4.0; weighted GEE models were implemented using SAS V9.4.

### Ethics approval

The EPICOV study received approval from an ethics committee (Comité de Protection des Personnes Sud Méditerranée III 2020-A01191-38) and from France’s National Data Protection Agency (Commission Nationale Informatique et Libertés, CNIL, MLD/MFI/AR205138).

## Results

Across the three study waves, the weighted prevalence of depression was respectively 17.1%, 14.9% and 13.8%. As shown in **Table 1**, 45.0% of young adults participating in the study were employed, 40.6% were studying, 9.2% were unemployed and 5.2% were out of the labor force. 59.5% of participants were aged 18-24 years, and 49.2% were women. A majority resided in urban areas (39.2% in a town >100,000 residents and 21.3% in the Greater Paris area). Most participants had a suitable (38.4%) or tight (32.4%) financial situation at study baseline and 17.4% reported income below poverty level. 19.7% of participants reported living alone, 9.1% had children, 41.0% were in a relationship and at study baseline a majority reported leaving their home at least once a week (42.9% 2-5 times and 23.2% >5 times). In terms of health, 12.5% of participants reported having a chronic disease and respectively 18.0%, 26.5% and 15.6% reported having experienced symptoms of COVID-19 in each of the three study waves.

**Table 1.** Descriptive characteristics of young adults (18-30 years) participating in the nationally representative EPICOV cohort study. France, 2020-2021, n=2217, weighted % and bivariate Generalized Estimating Equations (GEE) regression models testing associations with depression.

|  | **N**<br>**(%)** | **Depression**<br>**OR; 95% CI** |
| --- | --- | --- |
| **Labor market position:** Employed | 983 (45.0) | Ref |
| Student | 928 (40.6) | **1.76 [ 1.48 – 2.08]** |
| Unemployed | 194 (9.2) | **2.05 [1.58 - 2.65]** |
| Out of the labor force | 112 (5.2) | **1.77 [1.24 – 2.51]** |
| **Socio-demographic characteristics** |  |  |
| **Sex:** Male | 1013 (50.8) | Ref |
| Female | 1204 (49.2) | **1.59 [1.28 – 1.99]** |
| **Age (years):** 18-24 | 1347 (59.5) | Ref |
| 25-30 | 870 (40.5) | **0.64 [0.51 - 0.82]** |
| **Urbanicity:** Rural area | 376 (16.0) | Ref |
| <100 000 residents | 530 (23.5) | **1.38 [1.04 - 1.82]** |
| ≥ 100 000 residents | 785 (39.2) | **1.54 [1.20 - 1.98]** |
| Paris and suburbs | 393 (21.3) | **1.59 [1.21 - 2.09]** |
| **Financial situation:** Comfortable | 399 (17.4) | Ref |
| Suitable | 880 (38.4) | **1.32 [1.02 - 1.69]** |
| Tight | 697 (32.4) | **2.01 [1.56 – 2.59]** |
| Difficult | 234 (11.8) | **3.90 [1.40 - 2.80]** |
| **Lone living:** No | 1716 (80.4) | Ref |
| Yes | 416 (19.7) | **1.38 [1.15 - 1.65]** |
| **Has children:** No | 2002 (90.9) | Ref |
| Yes | 215 (9.1) | **0.41 [0.27 - 0.64]** |
| **In a relationship:** No | 714 (41.0) | Ref |
| Yes | 1089 (59.0) | **0.76 [0.63 – 0.92]** |
| **Outings in preceding week:** <once | 757 (33.9) | Ref |
| 2-5 times | 963 (42.9) | **0.77 [0.64 - 0.94]** |
| > 5 times | 497 (23.2) | **0.54 [0.44 - 0.66]** |
| **Health characteristics** |  |  |
| **Chronic disease:** No | 1932 (87.5) | Ref |
| Yes | 285 (12.5) | **1.66 [1.27 - 2.18]** |
| **COVID-19 Symptoms:** No | 1806 (82.0) | Ref |
| Yes | 411 (18.0) | **1.84 [1.54 – 2.20]** |

**Table 1** also shows bivariate associations between labor market participation, covariates, and depression across follow-up.

In a multivariate weighted statistical model controlling simultaneously for all covariates (**Table 2**), compared to participants who were employed the odds of depression were elevated among those who were students (OR: 1.29, 95% CI 1.05-1.60) and among those who were unemployed (OR: 1.50, 95% CI 1.13-1.99). Other factors associated with depression were being a woman (vs. man: OR: 1.58, 95% CI 1.34-1.86), being 25-30 years (vs. 18-24 years: OR: 0.80, 95% CI 0.65-0.98), living in an rural area (vs <100 000 residents: OR: 1.31, 95% CI 0.97-1.74; >=100 000 residents: OR: 1.30, 95% CI 1.01-1.69, Greater Paris area: OR: 1.51, 95% CI 1.13-2.01), financial situation (vs. comfortable respectively: suitable: OR: 1.38, 95% CI 1.07-1.79; tight: OR: 2.04, 95% CI 1.57-2.64 and difficult OR: 3.87, 95% CI 2.86-5.24), lone living (vs. living with somebody: OR: 1.31, 95% CI 1.08-1.59), having children (vs. no: OR: 0.42, 95% CI 0.28-0.64); outings in the preceding week (vs. <once, respectively: 2-5 times: OR: 0.80, 95% CI 0.64-0.98 and >5 times: OR: 0.62, 95% CI 0.49-0.79), having a chronic disease (vs. no: OR: 1.74, 95% CI 1.42-2.13) and experience of COVID-19 symptoms (vs. no: OR: 2.11, 95% CI 1.77-2.51).

**Table 2.** Association between labor market position and covariates and young people’s (18-30 years) symptoms of depression. Nationally representative EPICOV cohort study, France, 2020-2021, n=2217, multivariate Generalized Estimating Equations (GEE) regression models, weighted OR, 95% CI.

|  |  | OR | 95% CI |
| --- | --- | --- | --- |
| **Labor market participation** | Employed | Ref |  |
|  | Student | **1.29** | **[1.05 - 1.60]** |
|  | Unemployed | **1.50** | **[1.13 - 1.99]** |
|  | Out of the labor force | 1.42 | [0.98 - 2.08] |
| **Socio-demographic characteristics** |  |  |  |
| **Study wave** | 1 | Ref |  |
|  | 2 | 0.90 | [0.74 - 1.09] |
|  | 3 | 0.98 | [0.78 - 1.24] |
| **Sex** | Male | Ref |  |
|  | Female | **1.58** | **[1.34 - 1.86]** |
| **Age (years)** | 18-24 | Ref |  |
|  | 25-30 | **0.80** | **[0.65 - 0.98]** |
| **Urbanicity** | Rural area | Ref |  |
|  | <100 000 residents | 1.31 | [0.97 - 1.74] |
|  | >= 100 000 residents | **1.30** | **[1.01 - 1.69]** |
|  | Paris and suburbs | **1.51** | **[1.13 - 2.01]** |
| **Financial situation** | Comfortable | Ref |  |
|  | Suitable | **1.38** | **[1.07 - 1.79]** |
|  | Tight | **2.04** | **[1.57 - 2.64]** |
|  | Difficult | **3.87** | **[2.93 - 5.33]** |
| **Lone living** | No | Ref |  |
|  | Yes | **1.31** | **[1.08 - 1.59]** |
| **Has children** | No | Ref |  |
|  | Yes | **0.42** | **[0.28 - 0.64]** |
| **In a relationship** | No | Ref |  |
|  | Yes | 0.85 | [0.69 - 1.05] |
| **Outings in preceding week** | < once |  |  |
|  | 2-5 times | **0.80** | **[0.65 - 0.98]** |
|  | > 5 times | **0.62** | **[0.49 - 0.79]** |
| **Health characteristics** |  |  |  |
| **Chronic disease** | No | Ref |  |
|  | Yes | **1.74** | **[1.42 - 2.13]** |
| **Covid-19 symptoms** | No | Ref |  |
|  | Yes | **2.11** | **[1.77 - 2.51]** |

Stratifying the study population by age (**Table S1**), we found that among participants aged 18-24 years, compared to those who were employed, the odds of depression were elevated among students (multivariate OR: 1.33, 95% CI 1.03-1.70) and participants out of the labor force (multivariate OR: 1.71, 95% CI 1.04-2.82). Among participants who were unemployed the likelihood of depression was elevated but the OR did not reach statistical significance (multivariate OR: 1.41, 95% CI 0.96-2.09). In the 25-30 years age group, compared to participants who were employed, we found elevated odds of depression among those who were unemployed (multivariate OR: 1.78, 95% CI 1.17-2.71).

Stratifying the study population by sex (**Table S2**) we found no significant differences in the relationships between labor market participation and depression in men and women (compared to participants who were employed, multivariate ORs associated with being a student: men: 1.33, 95% CI 1.01-1.76; women: 1.19, 95% CI 0.85-1.67, multivariate Ors associated with being unemployed: men: 1.60, 95% CI 1.04-2.45; women: 1.47, 95% CI 1.01-2.15).

## Discussion

In a longitudinal nationally representative study of 18-30-year-olds, we found that labor market participation is associated with depression during the course of the COVID-19 epidemic in France. Young adults are a known high-risk group in terms of mental health; our study indicates that students and persons who are unemployed are especially vulnerable.

These results are robust to control for other characteristics associated with depression, highlighting the psychological needs of youths who are outside of the labor force and who should be the focus of psychological screening and intervention, particularly in the context of the COVID-19 pandemic.

### Limitations and strengths

Our study has limitations which need to be acknowledged prior to interpreting the data. First, the EPICOV study started in the Spring of 2020 and we have only retrospective data on participants’ history of pre-existing mental health problems. Nevertheless, we were able to exclude from the statistical analyses participants who had the most severe forms of psychopathology. While future studies should examine prospective changes in mental health among cohorts of young people followed since prior to the COVID-19 pandemic, our results do not appear to be due to pre-existing severe mental health difficulties. Second, the EPICOV cohort suffered attrition, with participants belonging to socioeconomically disadvantaged groups least likely to participate over the long term (17). This lead us to conduct repeated measures analyses, which make it possible to maximize the use of information provided by study participants and limit possible biases due to attrition. Moreover, our statistical analyses are weighted to render the study sample representative of young adults in France. Third, in the EPICOV cohort, depression was ascertained with the PHQ-9 questionnaire, which is self-reported. While clinical diagnoses may yield a more accurate picture of severe depression, the PHQ-9 has satisfactory validity vs. clinical diagnosis (18) and we specifically studied a level of symptoms consistent with moderate to severe depression.

Our study also has strengths which we would like to highlight. First, EPICOV is a nationally-representative sample of the population residing in France and the results are generalizable to the whole country (15). Second, to our knowledge, ours is one of few investigations in the context of the COVID-19 pandemic to directly compare young people in different labor market positions, including those who are out of employment and who are generally more difficult to include in health surveys and cohort studies. Third, EPICOV participants were followed-up from the Spring 2020 to the Fall 2021, making it possible to study participants’ mental health through different phases of the COVID-19 pandemic. Fourth, the use of online as well as telephone interviews and multiple call-backs helped include participants more reluctant to take part in health surveys in the study sample.

### Mental health of students

Our results are in line with other studies which found elevated levels of depression and anxiety among university students (3-5, 21). First of all, students are vulnerable because of high levels of financial difficulties as well social isolation, which increased during the course of the epidemic (21, 22). Additionally, there is evidence that students have experienced specific issues during the course of the COVID-19 pandemic, such as worries related to their studies, difficulties finding motivation to study, missed learning opportunities (23) as well as increased conflicts with others and limited ability to study (24), which could fuel fears about their future and negative mood. There is suggestion that distance learning is especially complex for students from disadvantaged backgrounds and negatively contributes to their stress levels (25). Additionally, students are one of the groups – along with adolescents, in which use of screen-based media and devices increased particularly during the course of the pandemic (22). While this helped them stay in contact with friends and family and pursue coursework, inability to socialize and meet others probably hampered the ability to share worries and anxieties and seek social support.

### Mental health of young adults who are unemployed

Young adults are especially likely to experience job instability which we found to be associated with depression, consistently with other studies (6, 7, 26). Importantly, while persons who experience mental health difficulties may be especially likely to become unemployed (27), there is also evidence that the experience of unemployment predicts later symptoms of depression and anxiety (28). The COVID-19 epidemic, which had a major impact on employment irrespective of individuals’ characteristics, is an interesting set-up to examine the relationship between unemployment and mental health, as pre-existing psychological difficulties are likely to play a lesser role than prior to the pandemic. Evidence showing that employment loss during the course of the sanitary crisis predicts psychological difficulties suggests that labor market characteristics could directly impact individuals’ mental health (10). Mechanisms that could contribute to elevated levels of depression among young adults who are unemployed include financial difficulties, stress associated with uncertainty about labor market prospects, as well as social isolation (8, 10). One of the issues that remains unresolved at this point is whether depression which occurred among young adults during the COVID-19 pandemic will persist or predict other mental health difficulties over the longer-term.

### Interventions promoting mental health in young people

The COVID-19 pandemic which had massive effects on population mental health, highlighted the need to test and make available interventions to prevent and treat persons who experience psychological distress and depression. Young adults have elevated risks of psychological distress, yet they are also one of the groups with lowest levels of access to mental healthcare (29). A recent umbrella review reported that psychosocial interventions as well as combined psychological and educational interventions lead by general practitioners are effective in preventing depression among youths (30). In addition, there is evidence that psychological counselling provided in the university context can effectively alleviate symptoms of depression among students (31). Moreover, mobile apps addressing signs of stress and psychological distress, which are effective and cheap, seem particularly well-suited to young adults who widely use screen-based media (32, 33). Mobile-based interventions and telemedicine may also be effective to address the mental health needs of young adults who are unemployed, although it may be more difficult to identify and target them. Additionally, efforts aiming to favor employment among young adults contribute to good mental health (34).

### Conclusions

Young adults experience high levels of depression in the context of the COVID-19 pandemic, students and the unemployed being at higher risk than young adults who are employed. Our findings imply that young adults who are in training or looking for work should be the focus of attention in terms of prevention and mental health treatment. In particular, efforts should be deployed to improve access to effective psychosocial interventions and mental health treatment for persons who are unemployed. Moreover, strategies that enhance young people’s chances of finding employment could have benefits in terms of mental health.

## Data Availability

All data produced in the present study are available upon reasonable request to the authors.

## Acknowledgements

The authors want to thank EPICOV study participants and members of the EPICOV study group, who all had a significant contribution to this study and made it possible.

## Data sharing

Anonymous aggregated data for the first wave of the EPICOV study are available online. The EPICOV dataset is available for research purposes concerning the first round, and will be available by the end of March 2022 for the second wave for research purpose on CASD (https://www.casd.eu/), after approval from French Ethics and Regulatory Committee (Comité du Secret Statistique, CESREES and CNIL). Access to anonymized individual data may become available before the expected period on request from the corresponding author, conditional on approval from an ethical and regulatory body.

