## Supplementary Tables for "Labor market participation and depression during the COVID-19 epidemic among young adults (18 to 30 years): a nationally representative study in France"

**Table S1. Association between labor market position and covariates and young people’s symptoms of depression by age. Nationally representative EPICOV cohort study, France, 2020-2021, multivariate Generalized Estimating Equations (GEE) regression models, weighted OR, 95% CI, p-value.**

|  |  | **18-24 years** | | **25-30 years** | |
| --- | --- | --- | --- | --- | --- |
|  |  | OR | 95% CI | OR | 95% CI |
| **Labor market participation** | Employed | Ref |  | Ref |  |
|  | Student | **1.33** | **[1.03 - 1.70]** | 1.31 | [0.79 - 2.16] |
|  | Unemployed | 1.41 | [0.96 - 2.09] | **1.78** | **[1.17 - 2.71]** |
|  | Out of the labor force | **1.72** | **[1.04 - 2.82]** | 1.00 | [0.53 - 1.87] |
| **Socio-demographic characteristics** | | | | | |
| **Study wave** | 1 | Ref |  | Ref |  |
|  | 2 | 1.04 | [0.82 - 1.31] | **0.67** | **[0.47 - 0.95]** |
|  | 3 | 1.05 | [0.79 - 1.41] | 0.87 | [0.59 - 1.29] |
| **Sex** | Male | Ref |  | Ref |  |
|  | Female | **1.61** | **[1.31 - 1.97]** | **1.55** | **[1.16 - 2.07]** |
| **Urbanicity** | Rural area | Ref |  | Ref |  |
|  | <100 000 residents | 1.33 | [0.91 - 1.94] | 1.28 | [0.79 - 2.09] |
|  | >= 100 000 residents | 1.36 | [0.96 - 1.91] | 1.25 | [0.80 - 1.95] |
|  | Paris and suburbs | **1.67** | **[1.15 - 2.43]** | 1.33 | [0.83 - 2.13] |
| **Financial situation** | Comfortable | Ref |  | Ref |  |
|  | Suitable | 1.33 | [0.97 - 1.81] | 1.50 | [0.94 - 2.39] |
|  | Tight | **1.91** | **[1.40 - 2.63]** | **2.27** | **[1.42 - 3.63]** |
|  | Difficult | **3.93** | **[2.72 - 5.68]** | **3.41** | **[1.99 - 5.83]** |
| **Lone living** | No | Ref |  | Ref |  |
|  | Yes | 1.12 | [0.87 - 1.47] | **1.65** | **[1.17 - 2.33]** |
| **Has children** | No | Ref |  | Ref |  |
|  | Yes | 0.77 | [0.36 - 1.64] | 0.42 | [0.26 - 1.68] |
| **In a relationship** | No | Ref |  | Ref |  |
|  | Yes | 0.99 | [0.76 - 1.28] | 0.66 | [0.47 - 0.94] |
| **Outings in preceding week** | < once | Ref |  | Ref |  |
|  | 2 - 5 times | **0.72** | **[0.56 - 0.93]** | 0.95 | [0.64 - 1.42] |
|  | > 5 times | **0.52** | **[0.38 - 0.69]** | 0.88 | [0.57 - 1.35] |
| **Health characteristics** |  |  |  |  |  |
| **Chronic disease** | No | Ref |  | Ref |  |
|  | Yes | **1.36** | **[1.05 - 1.77]** | **2.57** | **[1.42 - 3.63]** |
| **Covid-19 symptoms** | No |  |  |  |  |
|  | Yes | **2.05** | **[1.65 - 2.55]** | **2.22** | **[1.65 - 3.00]** |

**Table S2: Association between labor market position and covariates and young people’s symptoms of depression by sex. Nationally representative EPICOV cohort study, France, 2020-2021, multivariate Generalized Estimating Equations (GEE) regression models, weighted OR, 95% CI, p-value.**

|  |  | **Male** | | **Female** | |
| --- | --- | --- | --- | --- | --- |
|  |  | OR | 95% CI | OR | 95% CI |
| **Labor market participation** | Employed | Ref |  | Ref |  |
|  | Student | 1.19 | [0.85 - 1.67] | **1.33** | **[1.01 - 1.76]** |
|  | Unemployed | **1.60** | **[1.04 - 2.45]** | **1.47** | **[1.01 - 2.15]** |
|  | Out of the labor force | 1.44 | [0.80 - 2.59] | 1.52 | [0.92 - 2.51] |
| **Socio-demographic characteristics** | | | | | |
| **Study wave** | 1 | Ref |  | Ref |  |
|  | 2 | 1.14 | [0.85 - 1.54] | **0.73** | **[0.57 - 0.95]** |
|  | 3 | **1.46** | **[1.02 - 2.09]** | **0.73** | **[0.53 - 0.99]** |
| **Age (years)** | 18-24 | Ref |  | Ref |  |
|  | 25-30 | 0.79 | [0.57 - 1.09] | 0.78 | [0.60 - 1.03] |
| **Urbanicity** | Rural area | Ref |  | Ref |  |
|  | <100 000 residents | **1.74** | **[1.06 - 2.84]** | 1.12 | [0.78 - 1.60] |
|  | >= 100 000 residents | **1.84** | **[1.16 - 2.91]** | 1.07 | [0.77 - 1.48] |
|  | Paris and suburbs | **1.75** | **[1.07 - 2.87]** | **1.45** | **[1.01 - 2.08]** |
| **Financial situation** | Comfortable | Ref |  | Ref |  |
|  | Suitable | **1.70** | **[1.12 - 2.57]** | 1.19 | [0.85 - 1.66] |
|  | Tight | **2.35** | **[1.54 - 3.59]** | **1.87** | **[1.33 - 2.61]** |
|  | Difficult | **4.93** | **[3.06 - 7.95]** | **3.39** | **[2.29 - 5.01]** |
| **Lone living** | No | Ref |  | Ref |  |
|  | Yes | 1.34 | [0.97 - 1.84] | 1.26 | [0.97 - 1.63] |
| **Has children** | No | Ref |  | Ref |  |
|  | Yes | 0.49 | [0.22 - 1.07] | **0.42** | **[0.26 - 1.67]** |
| **In a relationship** | No | Ref |  | Ref |  |
|  | Yes | 1.06 | [0.76 - 1.48] | **0.71** | **[0.55 - 0.90]** |
| **Outings in preceding week** | < never | Ref |  | Ref |  |
|  | 2 - 5 times | **0.56** | **[0.41 - 0.78]** | 0.99 | [0.81 - 1.43] |
|  | > 5 times | **0.40** | **[0.28 - 0.57]** | 0.92 | [0.66 - 1.28] |
| **Health characteristics** | |  |  |  |  |
| **Chronic disease** | No | Ref |  | Ref |  |
|  | Yes | **2.14** | **[1.56 - 2.93]** | **1.53** | **[1.17 - 2.00]** |
| **Covid-19 symptoms** | No |  |  |  |  |
|  | Yes | **2.25** | **[1.71 - 2.96]** | **2.01** | **[1.60 - 2.53]** |
